# Systemic inflammation and increased tryptophan catabolism underpin depression, anxiety, and chronic fatigue symptoms after myocardial infarction: effects of revascularization

**DOI:** 10.64898/2025.12.23.25342889

**Authors:** Hussein Kadhem Al-Hakeim, Dhurgham Shihab Al-Hadrawi, Mengqi Niu, Michael Maes

## Abstract

**Background:** Patients with myocardial infarction (MI) often exhibit neuropsychiatric symptoms, but the underlying pathophysiological mechanisms remain elusive. This study examines the roles of the tryptophan catabolite (TRYCAT) pathway, systemic inflammation, and adverse metabolic remodeling in this comorbidity, both before and after percutaneous coronary intervention (PCI).

**Methods:** We assessed depression, anxiety, and chronic-fatigue syndrome (CFS)-like rating scales, blood levels of TRYCATs, inflammatory markers, and human fatty acid binding protein (h-FABP)4 in MI patients both before and after PCI and in healthy controls.

**Results:** MI patients exhibited significantly higher neuropsychiatric symptoms, a pro-inflammatory shift, TRYCAT pathway changes, and increased h-FABP4. The TRYCAT profile exhibited a bias towards neurotoxicity, characterized by elevated levels of 3-hydroxykynurenine and quinolinic acid and decreased kynurenic acid. Binary logistic regression revealed that a model using TRYCATs, immune data (e.g., neutrophil/lymphocyte ratio), and h-FABP4 distinguished post-MI patients from healthy controls with 92% cross-validated accuracy. A large part of the variance (around 60%) in the neuropsychiatric scores was explained by immune-metabolic data, higher neurotoxic and lower neuroprotective TRYCATs. PCI significantly improved neuropsychiatric symptoms, TRYCAT pathway-associated neuroprotection, the immune-inflammatory response, and h-FABP4.

**Conclusion:** MI is linked to increased TRYCAT-associated neurotoxicity and adverse inflammatory-metabolic remodeling in association with heightened depression, anxiety, and CFS-like symptoms. The reduction in immune activation, the normalization of neurotoxic TRYCAT synthesis, and reversal of adverse metabolic–inflammatory signaling following PCI may explain the observed improvement in neuropsychiatric symptoms following coronary revascularization.

## Introduction

Myocardial infarction (MI) is a leading cause of global morbidity and mortality (1). The principal pathogenesis often involves the rupture of an atherosclerotic plaque, leading to sudden thrombotic occlusion of a coronary artery and ensuing ischemia-induced necrosis of cardiac myocytes (2). The primary objective of treatment is to restore blood circulation, which can be achieved with percutaneous coronary intervention (PCI) (2). PCI is crucial in the treatment of acute MI as it safeguards the cardiac muscle and enhances clinical outcomes, including reduced angina and improved quality of life (3, 4). The lifesaving benefits of PCI may be undermined by ischemia-reperfusion injury, a situation where the restoration of blood flow initiates further damage, characterized by significant inflammatory and oxidative stress responses (5). The inflammatory response during MI and PCI is crucial for both immediate and prolonged cardiac remodeling (6, 7), underscoring the necessity for biomarkers that can precisely assess these injury and inflammation processes.

To precisely assess the dynamic damage process, biomarkers that distinguish specific pathophysiological pathways are essential. Fatty acid binding proteins (FABPs, including FABP3 and FABP4) have emerged as significant biomarkers for adverse cardiac remodeling. Ischemia rapidly induces the disintegration of the plasma membrane, resulting in the release of these low-molecular-weight cytoplasmic proteins (8). These proteins are sensitive early biomarkers of MI, defined as increased blood concentrations within 1-3 hours post-symptom onset (9). Therefore, assessing human fatty acid binding protein 4 (h-FABP4) levels before and during PCI may provide a dynamic representation of the severity of the first ischemic injury and perhaps quantify further damage resulting from ischemia-reperfusion injury (10).

The necrotic event and subsequent ischemia-reperfusion injury provoke a substantial systemic immune response, which induces considerable metabolic alterations. Recent research has emphasized the tryptophan catabolite (TRYCAT) pathway, the primary mechanism for tryptophan (Trp) breakdown, as a crucial link between immune function and metabolic flexibility (11). Pro-inflammatory cytokines, such as interferon-gamma (IFN-γ), significantly enhance the enzyme indoleamine 2,3-dioxygenase-1 (IDO1) in inflammatory circumstances. This alters the metabolic route for Trp from serotonin synthesis to kynurenine production, resulting in the formation of TRYCATs (12). TRYCATs are bioactive compounds that may affect the immune and nervous systems (13, 14). The overall equilibrium of these metabolites is crucial. The shift to neurotoxic TRYCATS, including 3-hydroxykynurenine (3-HK) and quinolinic acid (QA), is associated with heightened oxidative stress and pro-inflammatory reactions (15). In contrast, metabolites such as kynurenic acid (KA) possess antioxidant and anti-inflammatory properties (15). Modifications in this pathway have been associated with the exacerbation of atherosclerosis and heart failure (16, 17).

The TRYCAT pathway is significantly implicated in the pathophysiology of neuropsychiatric disorders (15, 18–20), including depression, anxiety, and chronic fatigue (21–23). Altered concentrations of TRYCATs, such as kynurenine (KYN) and KA, were observed in individuals with major depressive disorder (MDD) (24–26). This provides a plausible biochemical rationale for the increased prevalence of neuropsychiatric symptoms in MI patients (27, 28).

Thus, a significant pathophysiological link is shown between cardiomyocyte necrosis and this immune-metabolic response. The first events comprise a systemic immune-inflammatory response and adverse metabolic–inflammatory signaling, indicated by h-FABP4 release, and the consequent stimulation of the TRYCAT pathway (29). PCI, which may cause an additional inflammatory trigger, may further disrupt the delicate balance in the TRYCAT pathway (17, 30). Notwithstanding the documented alterations in the TRYCAT pathway among individuals with chronic cardiovascular issues (30, 31), our understanding remains insufficient regarding the temporal variations in the TRYCATs profile following the acute impacts of MI and PCI, as well as the correlation of these changes with those in h-FABP4, a marker of adverse inflammatory–metabolic response and remodeling.

Hence, this study aims to elucidate the longitudinal changes in TRYCATs, immune-inflammatory markers, and h-FABP4 in MI patients both before and after PCI. We aim to understand the relationship between the degree of myocardial injury and the TRYCAT pathway, chronic-fatigue syndrome (CFS)-like, anxiety, and depression symptoms.

## Subjects and Methods

### Subjects

Sixty coronary artery disease patients (male, aged 30-60 years) participated in this study. The patients were recruited from the Al-Najaf Center for Cardiac Surgery and Trans Catheter Therapy in Najaf, Governorate, Iraq, during the period from December 2024 to March 2025. The patients had acute MI, and they received standard medical therapy according to the European Society of Cardiology guidelines (32). Forty apparently healthy subjects were selected as a control group. They were recruited by word of mouth from the same catchment area. A senior psychiatrist used the Hamilton Depression Rating Scale (HAMD) to diagnose the score and severity of depression in all subjects. Exclusion criteria for patients and controls were renal or hepatic diseases, stroke, cancer, (auto)immune disorders, and neuroinflammatory conditions, and subjects with an albumin/creatinine ratio > 30 mg/g and serum C-reactive protein > 6 mg/dL, thereby excluding subjects with overt inflammation (33). Subjects with lifetime major depression, anxiety disorders, or CFS were excluded from participating. The body mass index (BMI) was calculated by dividing weight in kilograms by the square of height in meters. The Hamilton Anxiety Rating Scale (HAMA) was used to measure the severity of anxiety, and the Fibro-Fatigue scale to assess the severity of CFS-like symptoms. Based on these data, we computed an index of overall psychopathology as a z unit-based composite score summing up the z values of HAMD, HAMA, and Fibro-Fatigue (FF) scores.

Written informed consent was obtained from all subjects before participating in the study, which was approved by the IRB of the University of Kufa (487/2019) in compliance with the International Guidelines for Human Research protection as required by the Declaration of Helsinki.

### Assays

Each participant provided seven milliliters of blood, which was collected in two tubes. The subsequent step involved the rapid transfer of two milliliters of blood into tubes containing the anticoagulant dipotassium ethylenediaminetetraacetic acid (KLEDTA). Following the separation of components, the residual blood samples underwent centrifugation at 3000 rpm for 10 minutes to complete the clotting process. The tubes were subsequently stored at -80°C before analysis. Serum h-FABP4 and TRYCATs biomarkers (TRP, KA, KYN, 3-HK, and QA) were quantified by utilizing commercial ELISA sandwich kits provided by Melsin Medical Co., Jilin, China. All tests exhibit intra-assay coefficients of variation below 10%, indicating a high degree of accuracy. Colorimetric kits from Spectrum^®^ in Cairo, Egypt, were utilized to assess serum glucose, urea, and creatinine concentrations. Hematological parameters, such as white blood cell (WBC) count, were assessed using a five-part differential Mindray BC-5000 hematology analyzer (Shenzhen, China: Mindray Medical Electronics Co., Ltd.).

### Statistical Analysis

The distribution of the variables was examined utilizing the Kolmogorov-Smirnov test. Analysis of variance (ANOVA) was employed to compare continuous variables, while the Chi-square (χ^2^) test was utilized to assess associations among categorical variables. General linear model (GLM) analysis was used to compare biomarkers between the two groups, with age, sex, and BMI being adjusted when necessary. We performed a binary logistic regression analysis using the diagnosis of MI as the dependent variable (controls as the reference group) with biomarkers serving as explanatory variables. The odds ratio with 95% confidence intervals was calculated, along with the predictive accuracy, sensitivity, and specificity. Linear discriminant analysis (LDA) was used to evaluate the diagnostic performance of the biomarker profile by classifying subjects into their original group. Model performance and stability were assessed using the leave-one-out method. Multiple regression analysis was employed to identify the key biomarkers predicting the neuropsychiatric scale scores using both manual and stepwise automatic methods (p-to-entry of 0.05 and p-to-remove of 0.06). We calculated the standardized beta coefficients for each significant explanatory variable utilizing t-statistics with precise p-values, alongside the model F statistics and the total variance explained (R²), which served as an effect size. Additionally, the analysis was evaluated for homoscedasticity through the White and Breusch-Pagan tests, and for collinearity issues using variance inflation factor (VIF) and tolerance metrics. The Generalized Estimating Equations (GEE) model was used to analyze repeated measures data and included fixed effects of time (pre- and post-PCI). All statistical analyses were conducted using SPSS 30 (IBM-USA). Data transformations, such as log10, square root, rank, or Winsorization, were utilized, as necessary.

## Results

### Demographic and clinical characteristics

Demographic and clinical characteristics of the study participants are summarized in **Table 1**. MI patients and healthy controls were well-matched on all major demographic parameters, including age, sex, BMI, education, employment status, smoking status, and residence, with no statistically significant differences observed. MI patients exhibited a small but statistically significant reduction in peripheral oxygen saturation (SpOL) compared to controls. MI patients displayed higher scores on all measured psychopathology scales, including the FF, HAMA, and HAMD scores.

**Table 1.**
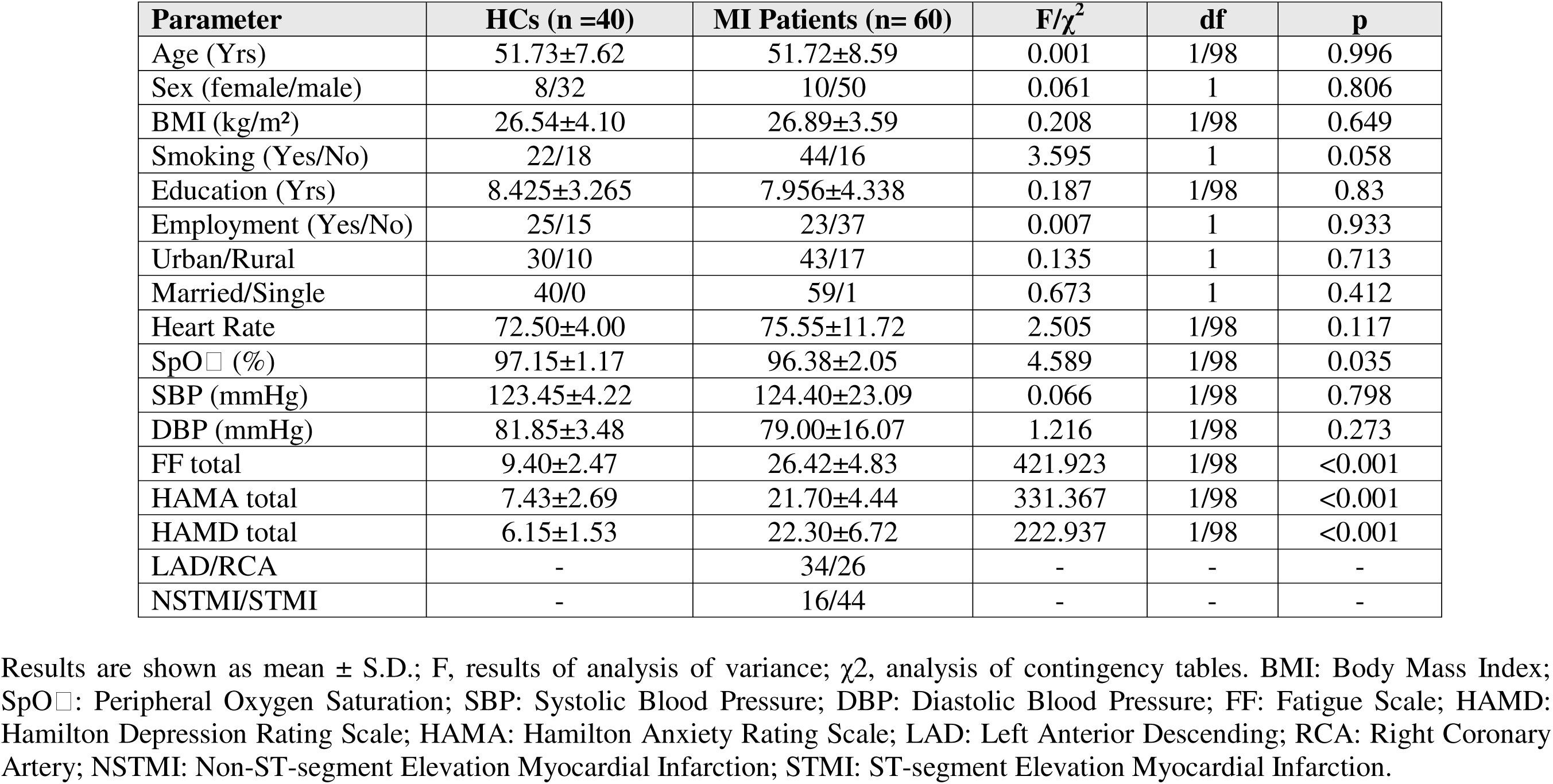
Demographics and clinical data between myocardial infarction (MI) patients and healthy controls (HCs)

| Parameter | HCs (n =40) | MI Patients (n= 60) | F/$\chi^2$ | df | p |
| --- | --- | --- | --- | --- | --- |
| Age (Yrs) | 51.73±7.62 | 51.72±8.59 | 0.001 | 1/98 | 0.996 |
| Sex (female/male) | 8/32 | 10/50 | 0.061 | 1 | 0.806 |
| BMI (kg/m<sup>2</sup>) | 26.54±4.10 | 26.89±3.59 | 0.208 | 1/98 | 0.649 |
| Smoking (Yes/No) | 22/18 | 44/16 | 3.595 | 1 | 0.058 |
| Education (Yrs) | 8.425±3.265 | 7.956±4.338 | 0.187 | 1/98 | 0.83 |
| Employment (Yes/No) | 25/15 | 23/37 | 0.007 | 1 | 0.933 |
| Urban/Rural | 30/10 | 43/17 | 0.135 | 1 | 0.713 |
| Married/Single | 40/0 | 59/1 | 0.673 | 1 | 0.412 |
| Heart Rate | 72.50±4.00 | 75.55±11.72 | 2.505 | 1/98 | 0.117 |
| SpO<sub>2</sub> (%) | 97.15±1.17 | 96.38±2.05 | 4.589 | 1/98 | 0.035 |
| SBP (mmHg) | 123.45±4.22 | 124.40±23.09 | 0.066 | 1/98 | 0.798 |
| DBP (mmHg) | 81.85±3.48 | 79.00±16.07 | 1.216 | 1/98 | 0.273 |
| FF total | 9.40±2.47 | 26.42±4.83 | 421.923 | 1/98 | <0.001 |
| HAMA total | 7.43±2.69 | 21.70±4.44 | 331.367 | 1/98 | <0.001 |
| HAMD total | 6.15±1.53 | 22.30±6.72 | 222.937 | 1/98 | <0.001 |
| LAD/RCA | - | 34/26 | - | - | - |
| NSTMI/STMI | - | 16/44 | - | - | - |

Results are shown as mean ± S.D.; F, results of analysis of variance; $\chi^2$ , analysis of contingency tables. BMI: Body Mass Index; SpO<sub>2</sub>: Peripheral Oxygen Saturation; SBP: Systolic Blood Pressure; DBP: Diastolic Blood Pressure; FF: Fatigue Scale; HAMD: Hamilton Depression Rating Scale; HAMA: Hamilton Anxiety Rating Scale; LAD: Left Anterior Descending; RCA: Right Coronary Artery; NSTMI: Non-ST-segment Elevation Myocardial Infarction; STMI: ST-segment Elevation Myocardial Infarction.

### Results of biomarkers

The estimated marginal means of biomarkers, adjusted for age, sex, and BMI, revealed profound and widespread differences in the TRYCAT and inflammatory profiles between MI patients and controls, as detailed in **Table 2**. MI patients exhibited significantly lower levels of TRP and the neuroprotective metabolite KA. Conversely, they had elevated levels of several neurotoxic metabolites, including KYN, 3-HK, and QA. This shift was further confirmed by a significantly higher KYN/TRP ratio, indicating increased IDO1 activity, and a lower KA/KYN ratio. MI patients exhibited a significantly elevated WBC count, increased neutrophils, a decrease in lymphocytes, and a higher neutrophil-to-lymphocyte ratio (NLR). Compared to controls, MI patients exhibited significantly elevated levels of glucose, serum creatinine, and h-FABP4.

**Table 2.** Differences in biomarkers between myocardial infarction (MI) patients and healthy controls (HC).

| Dependent Variable | HCs (n =40) | MI Patients (n= 60) | F | p |
| --- | --- | --- | --- | --- |
| Tryptophan (μM) | 61.91±2.22 | 38.78±1.81 | 65.09 | <0.001 |
| 3-OH-Kynurenine (nM) | 17.44±1.89 | 25.76±1.54 | 11.62 | <0.001 |
| Quinolinic acid (nM) | 217.39±20.36 | 321.59±16.62 | 15.70 | <0.001 |
| Kynurenic acid (nM) | 27.35±1.78 | 17.79±1.46 | 17.28 | <0.001 |
| Kynurenine (μM) | 1.93±0.18 | 3.06±0.15 | 22.39 | <0.001 |
| Anthranilic acid (nM) | 620.45±59.76 | 747.06±48.79 | 2.69 | 0.104 |
| KYN/TRP Ratio | 0.035±0.006 | 0.083±0.005 | 40.89 | <0.001 |
| KA/KYN Ratio | 17.59±1.98 | 11.29±1.62 | 6.06 | 0.016 |
| 3-HK/KYN Ratio | 11.14±1.51 | 10.79±1.24 | 0.03 | 0.860 |
| WBC /μL | 7.07±0.52 | 13.39±0.43 | 87.53 | <0.001 |
| Hemoglobin % | 14.20±0.20 | 14.70±0.16 | 3.75 | 0.056 |
| PCV % | 44.30±0.54 | 44.13±0.44 | 0.06 | 0.810 |
| Neutrophils % | 56.81±1.12 | 72.34±0.91 | 115.44 | <0.001 |
| Lymphocytes % | 32.31±0.98 | 20.87±0.80 | 81.65 | <0.001 |
| Neutrophil/Lymphocyte Ratio | 1.79±0.24 | 4.06±0.20 | 51.75 | <0.001 |
| Platelets /μL | 238.16±10.06 | 264.11±8.21 | 3.99 | 0.049 |
| Glucose (mM) | 5.61±0.25 | 7.76±0.20 | 45.97 | <0.001 |
| Blood Urea (mg/dl) | 31.82±1.36 | 34.13±1.11 | 1.71 | 0.194 |
| Serum Creatinine (mg/dl) | 0.67±0.48 | 3.09±0.39 | 15.52 | <0.001 |
| h-FABP4 (pg/ml) | 872.12±102.79 | 1284.42±83.91 | 9.65 | 0.002 |

All data are shown as marginal estimated mean ± SE; all df=1/98, all results of GLM analysis are adjusted for age and body mass index (residual values). KYN: Kynurenine; TRP: Tryptophan; KA: Kynurenic acid; 3-HK: 3-OH-Kynurenine; WBC: White Blood Cells; PCV: Packed Cell Volume; h-FABP4: human fatty acid-binding protein 4.

### Multivariate differences between MI and controls

To identify the most important biomarkers for distinguishing MI patients from controls, we performed binary logistic regression analyses, as shown in **Table 3**. Model #1, which uses all biomarkers as dependent variables, indicates that the 3-HK/KYN Ratio, WBC count, NLR, and h-FABP4 are significant predictors of MI relative to controls (Nagelkerke R² = 0.911). The model’s accuracy is 97.0% (sensitivity = 97.5%, specificity = 96.5%), with an AUC of 0.991. Other quality metrics of this model are the Gini index 0.982, maximum Kolmogorov-Smirnov statistic (Max K-S) 0.942, and overall model quality 0.98.

**Table 3.** Results of binary logistic regression analyses with myocardial infarction (MI) as the dependent variable and all biomarkers (Model #1) and only tryptophan catabolites (Model #2) as explanatory variables.

| Dichotomies | Explanatory variables | $\beta$ | SE | Wald | p | OR | 95% CI |
| --- | --- | --- | --- | --- | --- | --- | --- |
| #1. Patients vs. Controls | 3-HK/KYN | 1.533 | 0.660 | 5.387 | 0.020 | 4.630 | 1.269-16.893 |
|  | WBC | 4.898 | 1.574 | 9.679 | 0.002 | 134.048 | 6.125-2933.576 |
|  | Neutrophil/Lymphocyte | 3.181 | 0.981 | 10.519 | 0.001 | 24.059 | 3.520-164.434 |
|  | h-FABP4 | 1.282 | 0.649 | 3.906 | 0.048 | 3.604 | 1.011-12.852 |
| #2. Patients vs. Controls | Tryptophan | -3.448 | 0.943 | 13.364 | <0.001 | 0.032 | 0.005-0.202 |
|  | Quinolinic acid | 1.660 | 0.648 | 6.567 | 0.010 | 5.259 | 1.478-18.719 |
|  | Kynurenic acid | -1.511 | 0.666 | 5.143 | 0.023 | 0.221 | 0.060-0.815 |
|  | Kynurenine | 3.698 | 1.075 | 11.829 | <0.001 | 40.370 | 4.907-332.115 |
|  | KA/KYN | 1.375 | 0.736 | 3.489 | 0.062 | 3.956 | 0.934-16.753 |
|  | 3-HK/KYN | 2.157 | 0.821 | 6.906 | 0.009 | 8.646 | 1.730-43.201 |

All df=1. KYN: Kynurenine; KA: Kynurenic acid; 3-HK: 3-OH-Kynurenine; WBC: White Blood Cells; h-FABP4: human fatty acid-binding protein 4.

Model #2 included only the TRYCAT data and shows that QA, KA/KYN, 3-HK/KYN (positive correlations), TRP, and KA (negative correlation) were all significant predictors of MI. This model showed a Nagelkerke R² value of 0.983, a model accuracy of 93.0% (sensitivity = 87.5%, specificity = 96.7%), with an AUC of 0.973. Other quality metrics of this model are Gini index 0.945, Max K-S 0.858, and overall model quality 0.94. The leave-one-out cross-validated accuracy using LDA was 92.0%.

### Neuropsychiatric rating scale scores and biomarkers

Multiple regression analyses were performed to determine the extent to which serum biomarkers explain the variance in neuropsychiatric symptom scores, as shown in **Table 4**. Models (1–4) use all serum markers as explanatory variables, while Models (5–8) only use serum TRYCATs as the explanatory variable. Models 1-4 demonstrated that TRYCATs, WBC counts, and NLR were the most important predictors significantly contributing to explaining the variance in the FF, HAMA, HAMD, and total psychopathology score. Second, models 5-8 constructed solely with TRYCATs showed a consistent pattern emerging across all psychopathology measures; TRP was inversely correlated, whereas the neurotoxic metabolites QA and 3-HK were positively correlated with the outcome data. For example, TRYCAT alterations explained 46.1% of the variance in the total psychopathology score (Model 8).

**Table 4.** Results of multiple regression analyses with the neuropsychiatric rating scale scores as dependent variables, and serum biomarkers as explanatory variables.

| **Dependent Variables** | **Explanatory Variables** | **$\beta$** | **t** | **p** | **F model** | **df** | **p** | **R<sup>2</sup>** |
| --- | --- | --- | --- | --- | --- | --- | --- | --- |
| #1. FF total score | **Model** |  |  |  | 60.413 | 3/96 | <0.001 | 0.654 |
|  | WBC | 0.460 | 5.986 | <0.001 |  |  |  |  |
|  | Neutrophil/Lymphocyte | 0.386 | 4.899 | <0.001 |  |  |  |  |
|  | 3-OH-Kynurenine | 0.129 | 2.049 | 0.043 |  |  |  |  |
| #2. HAMA total score | **Model** |  |  |  | 87.031 | 2/97 | <0.001 | 0.642 |
|  | WBC | 0.453 | 5.827 | <0.001 |  |  |  |  |
|  | Neutrophil/Lymphocyte | 0.436 | 5.607 | <0.001 |  |  |  |  |
| #3. HAMD total score | **Model** |  |  |  | 27.030 | 5/94 | <0.001 | 0.590 |
|  | WBC | 0.452 | 5.030 | <0.001 |  |  |  |  |
|  | SBP | 0.198 | 2.961 | 0.004 |  |  |  |  |
|  | Neutrophil/Lymphocyte | 0.221 | 2.586 | 0.011 |  |  |  |  |
|  | Creatinine | 0.171 | 2.498 | 0.014 |  |  |  |  |
|  | Kynurenic acid | -0.152 | -2.171 | 0.032 |  |  |  |  |
| #4. Psychopathology score | **Model** |  |  |  | 106.064 | 2/97 | <0.001 | 0.686 |
|  | Fractional Rank of WBC | 0.518 | 7.107 | <0.001 |  |  |  |  |
|  | Fractional Rank of Neutrophil/Lymphocyte | 0.400 | 5.492 | <0.001 |  |  |  |  |
| #5. FF total score | **Model** |  |  |  | 19.596 | 4/95 | <0.001 | 0.452 |
|  | Tryptophan | -0.410 | -5.031 | <0.001 |  |  |  |  |
|  | Kynurenine | 0.381 | 4.238 | <0.001 |  |  |  |  |
|  | 3-HK/KYN | 0.294 | 3.378 | 0.001 |  |  |  |  |
|  | Quinolinic acid | 0.215 | 2.716 | 0.008 |  |  |  |  |
| #6. HAMA total score | **Model** |  |  |  | 14.769 | 4/95 | <0.001 | 0.383 |
|  | Tryptophan | -0.404 | -4.775 | <0.001 |  |  |  |  |
|  | Anthranilic acid | 0.250 | 3.075 | 0.003 |  |  |  |  |
|  | Quinolinic acid | 0.204 | 2.431 | 0.017 |  |  |  |  |
|  | 3-OH-Kynurenine | 0.203 | 2.413 | 0.018 |  |  |  |  |
| #7. HAMD total score | **Model** |  |  |  | 21.014 | 3/96 | <0.001 | 0.396 |
|  | Tryptophan | -0.401 | -4.816 | <0.001 |  |  |  |  |
|  | Quinolinic acid | 0.319 | 3.879 | <0.001 |  |  |  |  |
|  | 3-OH-Kynurenine | 0.246 | 3.002 | 0.003 |  |  |  |  |
| #8. Psychopathology score | **Model** |  |  |  | 20.297 | 4/95 | <0.001 | 0.461 |
|  | Tryptophan | -0.400 | -4.897 | <0.001 |  |  |  |  |
|  | Kynurenine | 0.178 | 2.201 | 0.030 |  |  |  |  |
|  | Quinolinic acid | 0.257 | 3.265 | 0.002 |  |  |  |  |
|  | 3-OH-Kynurenine | 0.243 | 3.064 | 0.003 |  |  |  |  |

Models 1-4 use all serum markers as explanatory variables, while Models 5-8 only use serum tryptophan catabolite as the explanatory variable. FF: Fibro-fatigue score; HAMD: Hamilton depression rating scale score; HAMA: Hamilton anxiety rating scale score; Psychopathology: an index based on the three former scores; KYN: Kynurenine; 3-HK: 3-OH-Kynurenine; WBC: White Blood Cells; SBP: Systolic Blood Pressure.

### Effects of PCI on biomarkers and clinical status

A GEE model, repeated measures design, was used to examine the effects of PCI on the clinical data and biomarkers. The results are detailed in **Table 5**. PCI treatment was associated with a significant decrease in all neuropsychiatric scores. Following PCI, levels of TRP increased, while KYN and 3-HK decreased. This was reflected in a substantial reduction in the KYN/TRP ratio, indicating decreased IDO1 activity, and a significant increase in the KA/KYN ratio. Levels of KA also rose significantly post-PCI. Furthermore, serum h-FABP4, creatinine, and WBC counts were significantly decreased after the procedure, alongside a small but significant improvement in SpOL.

**Table 5.** Results of GEE analysis, repeated measurements, examining the effects of intervention on biomarkers and clinical data.

| **Variables** | **Pre-PCI** | **Post-PCI** | **Wald** | **df** | **p** |
| --- | --- | --- | --- | --- | --- |
| Tryptophan (uM) | 38.86 (1.02) | 49.88 (1.88) | 26.280 | 1 | <0.001 |
| 3-OH-Kynurenine (nM) | 25.80(1.55) | 18.75(1.34) | 9.195 | 1 | 0.002 |
| Quinolinic acid (nM) | 314.64(18.95) | 313.42(23.38) | 0.002 | 1 | 0.969 |
| Kynurenic acid (nM) | 18.40(1.30) | 24.23(1.11) | 9.888 | 1 | 0.002 |
| Kynurenine (uM) | 3.15(0.18) | 2.51(0.13) | 6.683 | 1 | 0.010 |
| Anthranilic acid (nM) | 779.93(45.51) | 684.52(43.81) | 2.130 | 1 | 0.144 |
| KYN/TRP | 0.085(0.005) | 0.055(0.004) | 18.836 | 1 | <0.001 |
| KA/KYN | 9.30(0.86) | 12.50(0.75) | 12.880 | 1 | <0.001 |
| 3-HK/KYN | 10.79(1.28) | 8.23(0.69) | 2.422 | 1 | 0.120 |
| h-FABP4 (pg/ml) | 1306.92(111.68) | 893.60(51.30) | 9.016 | 1 | 0.003 |
| WBC /μL | 13.45(0.49) | 10.69(0.38) | 21.950 | 1 | <0.001 |
| Neutrophil/Lymphocyte Ratio | 4.19(0.27) | 3.76(0.17) | 1.561 | 1 | 0.211 |
| Serum Creatinine (mg/dl) | 3.03(0.51) | 0.86(0.06) | 18.081 | 1 | <0.001 |
| SpO<sub>2</sub> % | 96.39(0.26) | 96.89(0.17) | 6.585 | 1 | 0.010 |
| FF total score | 26.41(0.62) | 17.57(0.80) | 77.273 | 1 | <0.001 |
| HAMA total score | 21.69(0.57) | 12.74(0.42) | 130.062 | 1 | <0.001 |
| HAMD total score | 22.30(0.86) | 15.95(0.60) | 41.096 | 1 | <0.001 |
| Psychopathology (z score) | 1.94(0.21) | -1.94(0.18) | 181.547 | 1 | <0.001 |

KYN: Kynurenine; TRP: Tryptophan; KA: Kynurenic acid; 3-HK: 3-OH-Kynurenine; WBC: White Blood Cells; h-FABP4: heart-type fatty acid-binding protein 4. SpO<sub>2</sub>: Peripheral Oxygen Saturation; FF: Fibro-Fatigue Scale; HAMD: Hamilton Depression Rating Scale; HAMA: Hamilton Anxiety Rating Scale; Psychopathology: an index based on the three former scores.

GEE was used to evaluate the extent to which changes in biomarkers (from before to after the intervention) explained the changes in total psychopathology scores. We found that the overall psychopathology index was significant associated with the changes in h-FABP4 (F=8.94, p=0.003, B=+0.001, SE=0.0003), Trp (F=13.01, p<0.001, B=-0.049, SE=0.0136), 3-OHK (F=4.25, p=0.039, B=+0.044, SE=0.0215), KA (F=8.15, p=0.004, B=-0.060, SE=0.0210), KYN (F=7.48, p=0.006, B=+0.392, SE=0.1435), KYN/TRP (F=8.66, p=0.003, B=14.53, SE=4.939), KA/KYN (F=4.41, p=0.036, B=-0.054, SE=0.0259), but not with the other TRYCAT pathway markers. In addition, we could not detect any differences between the effects of LAD versus RCA on any of the biomarkers.

## Discussion

### Biomarkers of acute MI

The first major finding of this study shows that aberrations in the TRYCAT pathway, immune-inflammatory markers, and higher h-FABP4 are associated with MI. Our study shows that patients with acute MI exhibit a systemic immune-inflammatory response, characterized by increased WBC and NLR, along with elevated h-FABP4, indicating adverse metabolic–inflammatory signaling. The presence of an immune-inflammatory response and changes in FABPs have been reported previously (7, 8). The latter is considered to be an established biomarker of acute MI (9). Furthermore, the results of our logistic regression models show that the above assays are significant biomarkers of MI with an adequate cross-validated accuracy of 92%.

The significantly decreased levels of TRP, along with an elevated KYN/TRP ratio, might indicate increased inflammation-induced IDO1 activity. This enzyme is stimulated by pro-inflammatory cytokines such as IFN-γ (12, 34) and may lead to the accumulation of excessive neurotoxic metabolites (KYN, 3-HK, QA) and lowered levels of the neuroprotective KA in MI. These changes in neurotoxic versus neuroprotective TRYCATs profiles are known to be involved in other immune-inflammatory conditions and are associated with increased oxidative stress and immune activation (35). This "pathway shift" generally refers to a redistribution of TRP flux away from serotonin/melatonin synthesis toward a bias with increased 3-HK and QA production (13). Such metabolic reallocation is often interpreted as a signature of inflammation-driven metabolic reprogramming and may plausibly intersect with the pathophysiology of acute MI (35).

### The TRYCAT pathway and neuropsychiatric symptoms

The second major finding of this study is that MI is accompanied by a significant association between the immune-inflammatory response, including TRYCAT pathway activation, and neuropsychiatric symptoms, such as depression, anxiety, and CFS symptoms. Previously, it was described that TRYCAT pathway activation plays a role in cardiovascular disorders (CVD) and may explain the occurrence of mood symptoms as well as chronic fatigue in CVD (35). There is now evidence that the TRYCAT pathway is associated with severe subtypes of MDD (36).

We found that immune-inflammatory markers, like increases in WBC count and NLR, are strongly associated with the severity of neuropsychiatric symptoms in MI patients. This supports the neuroimmune, metabolic, and oxidative stress (NIMETOX) theory of depression (37) whereby systemic immune activation is considered to contribute to central neuroinflammation and aberrations in limbic-prefrontal cortex circuits, leading to affective symptoms. In addition, stimulation of IDO1 via immune activation may contribute to these central disorders through the production of neurotoxic TRYCATs and TRP depletion (15).

### PCI improves neuropsychiatric symptoms and biomarkers

The third major finding of this study is that PCI induced a rapid reduction in all neuropsychiatric rating scores in association with significant improvement of the biomarkers. Therefore, after treatment, TRP levels and KA/KYN ratio increased, whilst KYN, 3-HK, KYN/TRP, h-FAPB, and WBC numbers diminished. These findings provide a plausible explanation for the rapid relief of psychiatric symptoms often observed post-revascularization (38–40). It is well known that PCI reinstates coronary flow (41). Thus, by treating the ischemic injury, mitigating the immune-inflammatory response, and reversing the adverse metabolic-inflammatory signals, PCI might diminish IDO1 activation, thereby promoting a more favorable rebalancing of the TRYCAT pathway towards neuroprotective and less neurotoxic activities.

### Limitations

This study has certain limitations that should be acknowledged when interpreting the results. The study was conducted at a singular location, which restricts its broader applicability. These findings require validation across diverse cultures and nations. We evaluated the most significant TRYCATs. However, we did not quantify the activity or expression levels of enzymes involved in the TRYCAT pathway, such as IDO1, KAT, or KMO, although metabolite ratios, such as KYN/TRP as an indicator of IDO1 activity, serve as a widely accepted indirect measure of enzyme function. This is a case-control study; therefore, causality cannot be established.

## Conclusions

The current study demonstrates that acute MI is associated with neuropsychiatric symptoms, which are linked to activation of immune-inflammatory pathways, TRYCAT pathway engagement, and detrimental metabolic-inflammatory signaling. The reduction in immune activation and the normalization of IDO1-driven neurotoxic TRYCAT synthesis may explain the observed improvement in neuropsychiatric symptoms following revascularization. Our findings indicate that the TRYCAT pathway serves as a significant connection among cardiac injury, systemic immune activation, and neuropsychiatric symptoms.

## Data Availability

The database created during this investigation will be provided by the corresponding author (MM) upon a reasonable request once the authors have thoroughly used the data set.

## Ethics approval

This study was approved by the IRB of the University of Kufa (487/2019) in compliance with the International Guidelines for Human Research protection as required by the Declaration of Helsinki.

## Consent to participate

Before participating in this study, each subject provided written informed consent.

## Consent for publication

All authors have given their approval for this paper to be published.

## Author’s contributions

Hussein Kadhem Al-Hakeim: conceptualization, data curation, supervision and writing - original draft; Dhurgham Shihab Al-Hadrawi: data curation and writing - review & editing; Mengqi Niu: writing - review & editing; Michael Maes: conceptualization, formal analysis, supervision and writing - review & editing. All the contributing authors have participated in the preparation of the manuscript.

